# Automatic Responsiveness Testing in Epilepsy with Wearable Technology: The ARTiE Watch

**DOI:** 10.1101/2024.05.27.24307959

**Authors:** Lydia Wheeler, Vaclav Kremen, Cole Mersereau, Guillermo Ornelas, Taruna Yadav, Devon Cormier, Allyson Derry, Andrea Duque Lopez, Kevin McQuown, Vladimir Sladky, Christopher Benjamin, Joseph Giacino, START Project Collaborative Team, Gregory Worrell, Hal Blumenfeld

## Abstract

**Objective:** An accurate evaluation of behavioral responsiveness during and after seizures in people with epilepsy is critical for accurate diagnosis and management. Current methods for assessing behavioral responsiveness are characterized by substantial variation, subjectivity, and limited reliability and reproducibility in ambulatory and epilepsy monitoring unit (EMU) settings.

In this study, we aimed to develop and implement a novel mobile platform for deployment of automated responsiveness testing in epilepsy – the ARTiE Watch, to facilitate standardized, objective assessments of behavioral responsiveness during and after seizures.

**Methods:** We prospectively recruited patients admitted to the epilepsy monitoring units for diagnostic evaluation and long-term video-electroencephalographic monitoring at Mayo Clinic and Yale New Haven Hospital. Participants wore the ARTiE Watch, a smartwatch paired with custom smartphone software integrated with cloud infrastructure allowing for remote activation of standardized assessment on the participants’ smartwatch. The assessment consisted of 18 command prompts which test behavioral responsiveness across motor, language, and memory domains.

Upon visually identifying an electrographic seizure during EMU monitoring, the BrainRISE platform was used to deploy the ARTiE Watch behavioral testing sequence. Responsiveness scoring was conducted on smartwatch files.

**Results:** Eighteen of 56 participants had a total of 39 electrographic seizures assessed with the ARTiE Watch. The 18 subjects with ARTiE Watch-tested seizures had a total of 67 baseline (interictal) ARTiE Watch tests collected for analysis. The analysis showed distinct ARTiE Watch behavioral responsiveness phenotypes: (1) decreased responsiveness across all ARTiE Watch commands during seizure (ictal-postictal) periods compared (p<0.0001) to baseline, (2) decreased responsiveness in bilateral tonic-clonic seizures compared to baseline (p<0.0001) and compared (p<0.0001) to focal seizures, and (3) decreased responsiveness during focal impaired awareness seizures compared (p<0.0001) to baseline and compared (p<0.001) to focal aware seizures.

**Significance:** ARTiE Watch behavioral testing deployed utilizing a mobile cloud-based platform is feasible and can provide standardized, objective behavioral responsiveness assessments during seizures.

**Key points:**

- cloud-based platform was used to deliver ARTiE Watch testing during interictal (baseline), ictal, and post-ictal periods in epilepsy monitoring unit participants.
- Watch behavioral responsiveness testing shows different behavioral responsiveness phenotypes for focal aware seizures (FAS), focal impaired aware seizures (FIAS), and bilateral tonic-clonic seizures (BTC).
- Watch behavioral responsiveness testing deployed utilizing a cloud-based platform provides reproducible, standardized, objective behavioral assessments.

## 1 INTRODUCTION

Evaluation of behavioral responsiveness during (ictal), after (postictal), and between (interictal) seizures is critical for accurate diagnosis, classification, and management in people with epilepsy (PWE) (Hanrahan et al., 2021). The gold standard for definitive epilepsy diagnosis is video-electroencephalography (VEEG) monitoring to record the patients’ habitual, spontaneous seizures (Devinsky et al. 2018). The clinico-electrophysiologic correlation and interactive behavioral assessment by epilepsy monitoring unit (EMU) personnel can be performed during the ictal and postictal phases of seizures. Such assessments are interactive and consist of a series of commands used to assess language, orientation, memory, and motor function. These behavioral assessments help to inform classification of seizure type based on seizure semiology and focal deficits. The results of behavioral testing during seizures are important for diagnosis and treatment recommendations (Beniczky et al., 2016). However, there is currently substantial variation in behavioral testing, and it is currently largely limited to applications in hospitalized patients undergoing EMU evaluations (Hamandi et al., 2017). The lack of standardized behavioral assessments limits our ability to utilize the wealth of behavioral data to reveal distinct behavioral phenotypes (Hanrahan et al., 2021; Toulomes et al. 2016; Kinney, Kovac, Diehl 2019). Furthermore, a fundamental gap in clinical epileptology is the objective characterization of seizure semiology of seizures.

We aimed to address these clinical gaps with a cloud-based platform technology that integrates a smartwatch and smartphone to deliver standardized automated responsiveness testing in epilepsy (the ARTiE Watch). Here, we evaluate the ARTiE Watch behavioral responsiveness system in a cohort of participants with epilepsy in two level 4 epilepsy centers.

## 2 MATERIALS AND METHODS

### 2.1 Study design

Adult patients admitted to the Epilepsy Monitoring Unit (EMU) at Mayo Clinic Rochester, Minnesota and Yale New Haven Hospital, New Haven, Connecticut for diagnostic evaluation between May 2021 and July 2023 were identified. The inclusion criteria were: (1) age ≥ 18 years, (2) ability to perform behavioral tasks on a smartwatch at baseline, and (3) confirmed or clinical suspicion of epilepsy. This prospective cohort study was approved by the Mayo Clinic Institutional Review Board and Yale University Human Investigations Committee. Verbal and written informed consent were obtained from each participant.

### 2.2 Automatic Responsiveness Testing in Epilepsy (ARTiE) Watch

The automatic responsiveness testing in epilepsy (ARTiE) Watch is a semi-automated behavioral testing system that can be used in the clinical EMU and ambulatory settings. The main components in the current study include the (1) ARTiE Watch custom software application adapted to run on a smartphone and smartwatch and (2) BrainRISE a cloud-based platform for integration and remote activation of the ARTiE Watch behavioral assessment (Kremen et al. 2024, 2018). The bidirectional wireless connectivity between smartphone, smartwatch, and cloud environment enables remote initiation of ARTiE Watch testing and cloud-based centralized data storage. The BrainRISE platform provides a seamless ecosystem for storage, synchronization, viewing, annotation, and analysis of multi-modal data streams in ambulatory humans (Kremen et al. 2024, 2018). All data is synchronized and aggregated in a longitudinal record format and are mapped directly to annotated timestamps.

**Figure 1.**
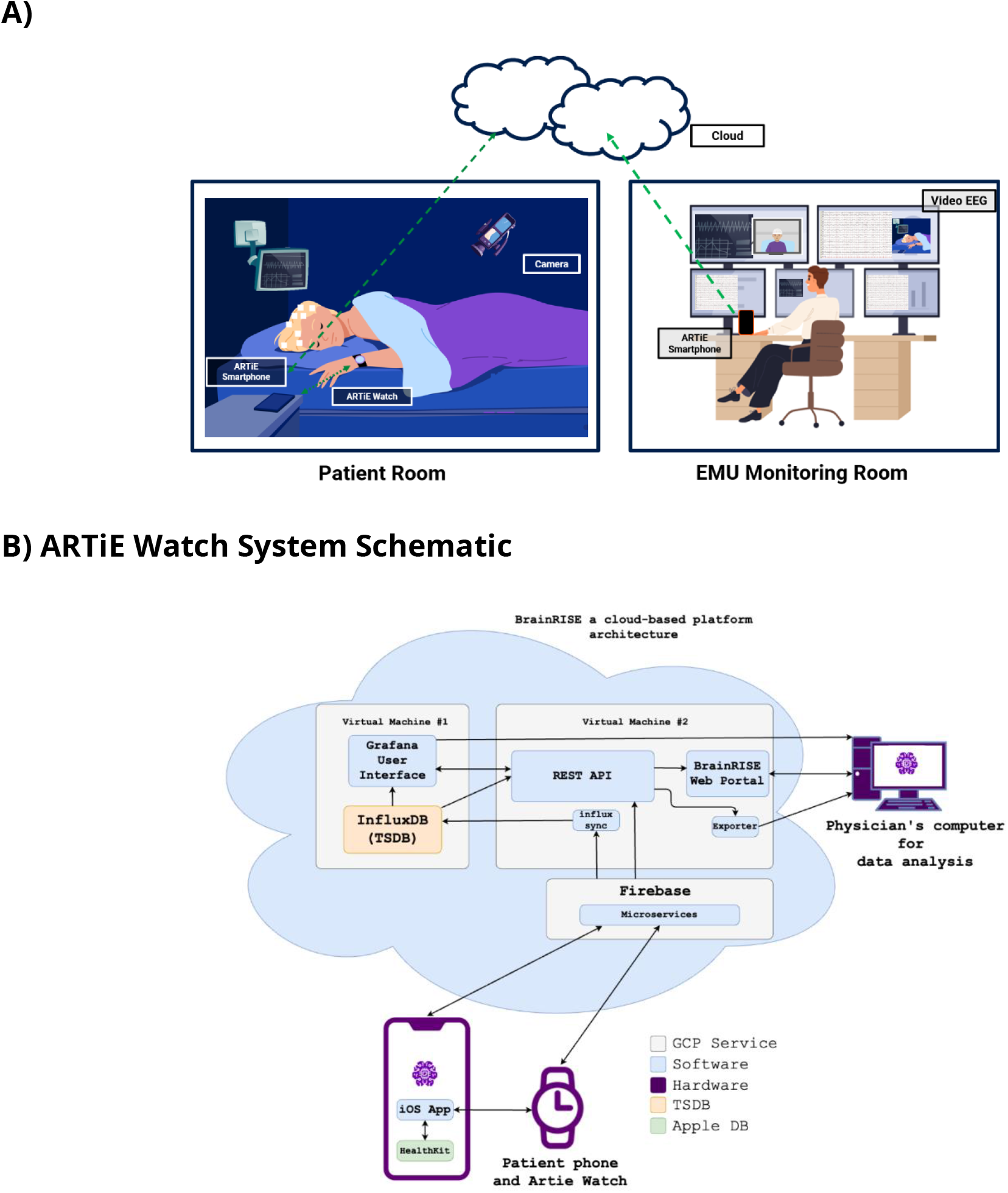
Schematic of Automatic responsiveness testing in epilepsy (ARTiE) Watch Behavioral Responsiveness System in seizures recorded in the EEG Epilepsy Monitoring Unit (EMU). (a) The platform for ARTiE Watch testing in the EMU utilizes a cloud-based application to trigger questionnaires, acquire patient responses, and synchronize behavioral and EEG data streams. Upon patient event button press or visual detection of an EEG seizure by EMU monitoring staff, the ARTiE Watch testing is remotely initiated on the patient’s smartwatch. (b) A simplified block diagram of the cloud-based and mobile platform using an ARTIE watch and mobile phone in the patient’s EMU room. Once the ARTIE Watch testing is started, the watch collects the data including recorded audio signals and tactile responses on the screen, and the data are then automatically pushed to the cloud database (Firebase). Cloud architecture automatically distributes the data and audio to the Influx database (InfluxDB – Time Series Database (TSDB)), which servers the data in the Grafana environment. The BrainRISE web portal then uses a Representational State Transfer (REST) Application Programming Interface (API) to receive and present the data to the end user (in this case, physician, or study team member) for data download, review and analysis. The mobile application provides an easily accessible interface for standardized behavioral testing during seizures while the cloud environment automatically receives and serves the data for subsequent analytics.

In the current study we manually triggered the mobile ARTiE Watch assessment upon visually identifying an electrographic seizure from the EMU monitoring station (Figure 1). The EMU monitoring technician used the BrainRISE application to launch a remote notification to the cloud server (wi-fi or cellular connectivity) which sends a push notification to the patient smartphone (wi-fi or cellular connectivity) and initiates the ARTiE Watch testing and recording sequence ((cellular connectivity or Bluetooth Low Energy (BLE)). The data collected from the ARTiE Watch is automatically synchronized with cloud notifications.

### 2.3 ARTiE Watch Behavioral Testing Sequence

The ARTiE Watch behavioral responsiveness testing sequence was modified from previously validated human-administered behavioral testing batteries such as the JFK Coma Recovery Scale-Revised (Giacino, Kalmar, and Whyte 2004), and the Responsiveness in Epilepsy Scale (Yang et al., 2012; Bauerschmidt et al., 2013; Cunningham et al., 2014). The initial implementation of automatic responsiveness testing in epilepsy (ARTiE) was on a tablet computer (Toulomes et al. 2016). The testing sequence was modified further for the present fully mobile platform. The current ARTiE Watch behavioral responsiveness testing sequence consists of 18 command prompts (Table 1; Figure 2; Supplementary Video 1). The ARTiE Watch test length is 6 minutes and 56 seconds, which is longer than the average seizure duration of 1-2 minutes, to allow for capture of ictal and postictal behavioral responsiveness (Afra et al., 2008; O’Kula et al., 2021).

**Figure 2.**
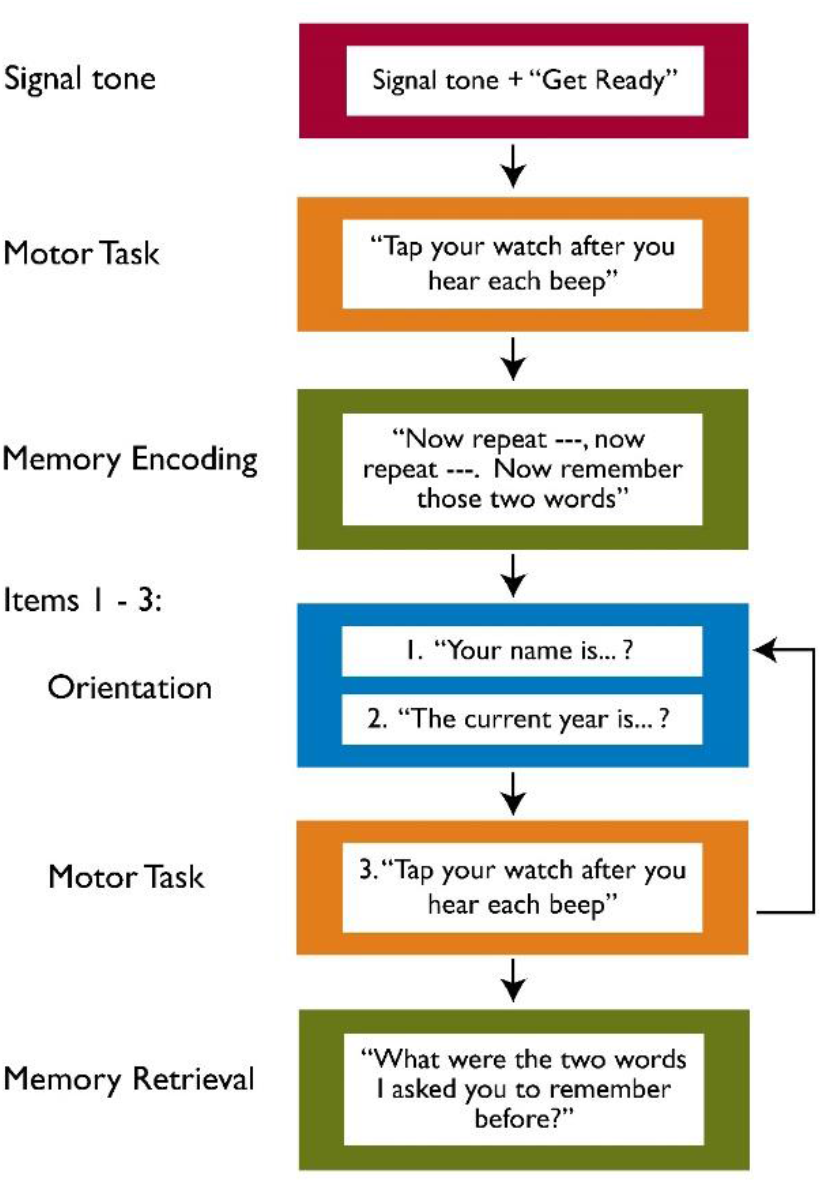
Schematic of Automatic responsiveness testing in epilepsy (ARTiE) Watch Behavioral Testing Sequence. Total duration of the ARTiE Watch behavioral testing from the start of the initial Signal Tone until the end of the Memory Retrieval question is 6 minutes 56 seconds, and consists of a total of 18 test items. Items 1-3 are repeated in a total of 5 cycles, with a pause in testing after each cycle.

At ARTiE Watch test onset, an auditory signal tone plays on the smartwatch to alert the participant that the ARTiE Watch behavioral testing sequence and audio recording is beginning on the smartwatch. The smartwatch asks the participant to complete a motor task (audio command “tap your watch after you hear each beep”) and then a memory encoding task (audio command to repeat and remember two words). The sequence then enters 5 loops of verbal orientation questions (asks participant to say their “name” and the “current year”) and motor tasks (“tap your watch after you hear each beep”). The final task is memory recall (“what were the two words I asked you to remember before?”) (Table 1).

### 2.4 ARTiE Watch Test Initiation and Behavioral Scoring

Enrolled patients were monitored by certified EEG monitoring technicians during the vEEG from the EMU monitoring room. Immediately upon visual detection of seizure onset (clinical or electrographic) or patient event button press, EMU staff remotely initiated the ARTiE Watch behavioral testing sequence on the patient’s smartwatch. The staff then went to the patient’s bedside. Interictal (baseline) tests were administered daily, during an hour without ictal activity or non-electrographic clinical events, to obtain ARTiE Watch baseline scores for each patient.

For scoring of ARTiE Watch behavioral testing, audio and behavioral files from the watch were transferred via the cloud to a local computer for analysis at each site. Timing of each test item relative to seizure onset and termination was determined based on simultaneous video/EEG. Each ARTiE test was reviewed and scored independently by two individuals at each site. An item score of 0 was assigned when there was no response, 3 was assigned when there was a normal response, and scores of 1 or 2 for intermediate levels of abnormal responses (Table 1). Audio files recorded by the ARTiE Watch during testing were used to score all items requiring a verbal response (Table 1). The Motor Task was scored automatically by the BrainRise software, with a correct response scored for any tap occurring after each beep onset and prior to onset of the subsequent beep. Two seconds were allowed for response to the last beep. When multiple taps occurred after a single beep only the first tap was scored.

### 2.5 Video-EEG (vEEG) Review and Clinical Seizure Classification

Continuous vEEG data (Natus Neuroworks, Natus, Pleasanton, CA) were acquired from scalp or intracranial electrodes in all patients. Two epileptologists visually reviewed the vEEG to: (1) identify and annotate EEG seizure onset and offset; (2) characterize behavior, seizure semiology, clinical onset and offset; (3) characterize seizure localization and classification according to ILAE guidelines (Fisher et al., 2017); and (4) identify the timing of ARTiE onset relative to vEEG seizure onset and offset. EMU clinical staff assessments were used to classify focal onset seizures as focal aware seizures (FAS), focal impaired awareness seizures (FIAS), and tonic-clonic (TC, including focal to bilateral tonic-clonic and generalized onset tonic-clonic seizures) (Fisher et al., 2017). Focal seizure onset locations were further classified by the clinical epileptology teams at each site using all available clinical information, including clinical history, video and scalp or intracranial EEG, neuroimaging, and surgical outcome if available.

### 2.6 Statistical analysis

Statistical analyses were performed using the R Project for Statistical Computing software package (https://www.r-project.org/). Nonparametric Kruskal-Wallis or Wilcoxon rank-sum tests were used for comparison of behavioral responsiveness. We combined ARTiE Watch scores into the following categories: Verbal Responsiveness, Motor Responsiveness, Memory Recall, and Overall Responsiveness (e.g. see Figure 3 B – E). The Verbal Responsiveness score includes the mean score of all language tasks (memory encoding, name, and year) excluding memory recall. The Memory Recall score is reported separately and includes the mean of the memory recall tasks. The Motor (nonverbal) Responsiveness score includes the mean score of motor tasks (“tap the watch after each beep”). The Overall Responsiveness score includes the mean score of all tasks (motor and verbal) excluding memory recall. Data are shown as mean ± SEM. All pairs were analyzed for significance, Bonferroni adjustments for multiple comparisons were performed, and statistical significance was set at *p* < 0.05 and is indicated as follows: ****p ≤ 0.0001, ***p ≤ 0.001, **p ≤ 0.01, *p < 0.05, ns – nonsignificant (p ≥ 0.05).

## 3 RESULTS

### 3.1 Participant Demographics

A total of 56 people epilepsy were enrolled in the ARTiE Watch EMU study across the two institutions. Of the 56 participants, 18 (32%) participants had a total of 39 electrographic seizures captured with ARTiE Watch behavioral responsiveness testing. Participant demographics are presented in Table 2, and were similar for the 56 total patients enrolled and for the 18 with seizures captured by the ARTiE Watch. Of the 56 total participants enrolled, twenty-nine (52%) participants were female and 27 (48%) were male. Fifty (89%) underwent scalp EEG and 6 (11%) underwent intracranial EEG. The mean age of participants was 34.2 (range 19-62) years. Forty-three (77%) were right-handed, 11 (20%) were left-handed, and 2 (3%) were ambidextrous. Forty-seven (84%) participants had drug resistant epilepsy and 3 (5%) had a history of status epilepticus. - The most common comorbid conditions were anxiety (41%), depression (38%), and migraine (36%).

**Table 2.**
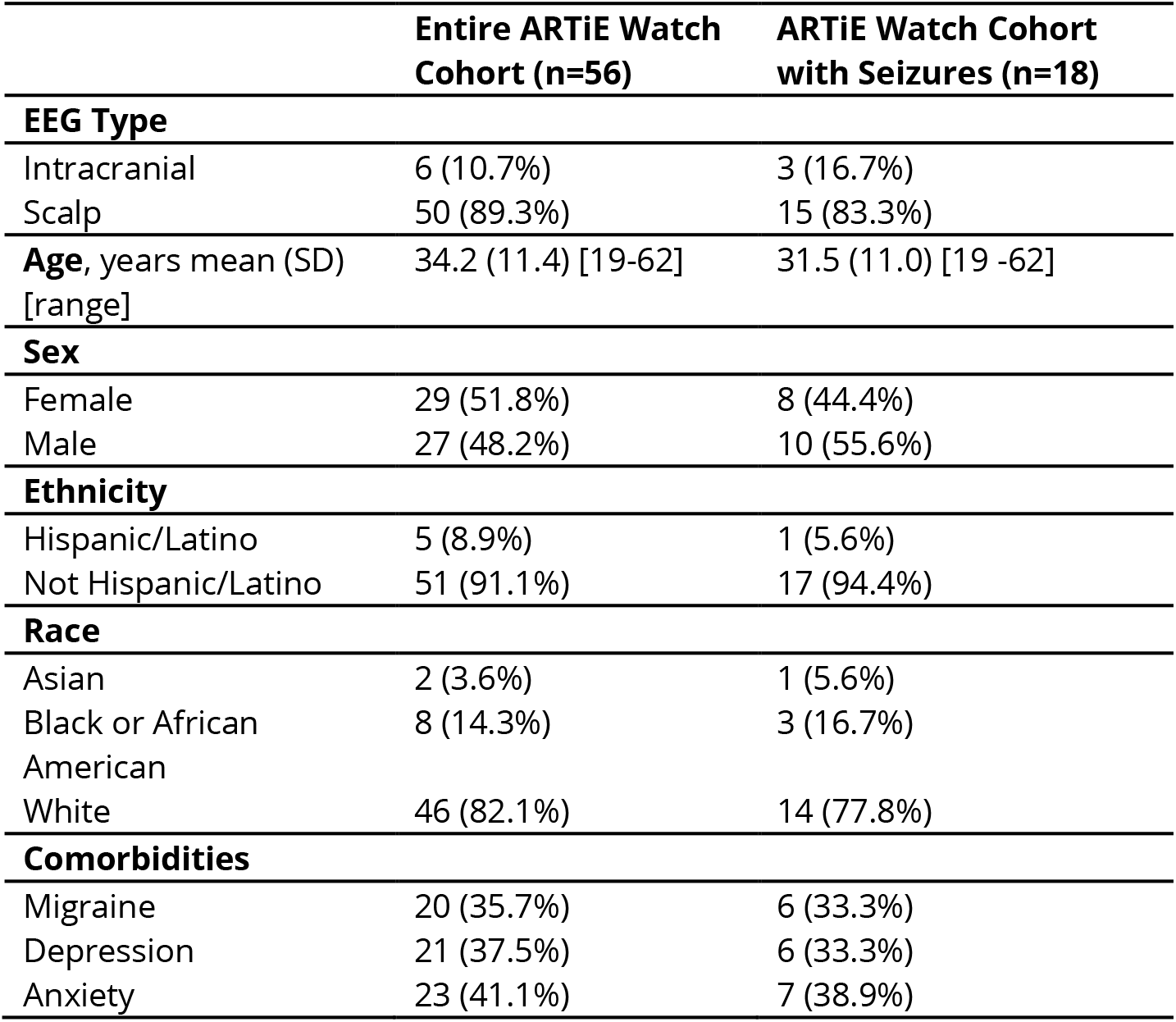
ARTiE Watch Behavioral Responsiveness Participant Characteristics.

|  | **Entire ARTiE Watch Cohort (n=56)** | **ARTiE Watch Cohort with Seizures (n=18)** |
| --- | --- | --- |
| **EEG Type** |  |  |
| Intracranial | 6 (10.7%) | 3 (16.7%) |
| Scalp | 50 (89.3%) | 15 (83.3%) |
| **Age**, years mean (SD) [range] | 34.2 (11.4) [19-62] | 31.5 (11.0) [19 -62] |
| **Sex** |  |  |
| Female | 29 (51.8%) | 8 (44.4%) |
| Male | 27 (48.2%) | 10 (55.6%) |
| **Ethnicity** |  |  |
| Hispanic/Latino | 5 (8.9%) | 1 (5.6%) |
| Not Hispanic/Latino | 51 (91.1%) | 17 (94.4%) |
| **Race** |  |  |
| Asian | 2 (3.6%) | 1 (5.6%) |
| Black or African American | 8 (14.3%) | 3 (16.7%) |
| White | 46 (82.1%) | 14 (77.8%) |
| **Comorbidities** |  |  |
| Migraine | 20 (35.7%) | 6 (33.3%) |
| Depression | 21 (37.5%) | 6 (33.3%) |
| Anxiety | 23 (41.1%) | 7 (38.9%) |

For the 18 (32%) participants included in the final analysis, 8 (44%) were female and 10 (56%) were male. Fifteen (83%) underwent scalp EEG and 3 (17%) underwent intracranial EEG. The mean age of the 18 participants was 31.5 (range 19-62) years. Twelve (67%) were right-handed and 6 (33%) were left-handed.

Clinical characteristics are presented in Table 3 for all 56 enrolled participants, as well as for the 18 patients with seizures tested with the ARTiE Watch. For the 18 patients included in the final analysis, the mean age of epilepsy onset was 19.8 (range 1.5 – 54) years, epilepsy duration was 11.2 (range 1-30) years, and monthly seizure frequency was 11.2 (range 0.17 – 60). Fifteen (83%) had drug resistant epilepsy, 3 (17%) had neuromodulation devices, and 14 (78%) took two or more antiseizure medications (ASMs).

**Table 3.** Clinical Characteristics of Epilepsy in ARTiE Watch Cohort.

|  | **Entire ARTiE Watch Cohort (n=56)** | **Cohort with Seizures (n=18)** |
| --- | --- | --- |
| **Age at Epilepsy Onset** mean (SD) range | 19.7 ± 12.3 [1.5-54] | 19.8 ± 13.8 [1.5-54] |
| **Epilepsy Duration** (years) | 15.2 (11.7) [1-45] | 11.2 (8.3) [1-30] |
| **Monthly Seizure Frequency** | 11.9 (18.1) [0.08 – 75] | 13.7 (16.5) [0.17 – 60] |
| **Drug Resistant Epilepsy** | 47 (83.9%) | 15 (83.3%) |
| **History of Status Epilepticus** | 3 (5.4%) | 0 (0%) |
| **Number of Antiseizure Medications (ASMs)** |  |  |
| One | 11 (19.6%) | 4 (22.2%) |
| Two | 23 (41.1%) | 7 (38.9%) |
| Three or More | 21 (37.5%) | 7 (38.9%) |
| **Epilepsy Treatment** |  |  |
| ASM | 35 (62.5%) | 13 (72.2%) |
| ASM + Neuromodulation Device | 8 (14.3%) | 3 (16.7%) |
| ASM + Prior Resection | 12 (21.4%) | 2 (11.1%) |
| **Epilepsy Etiology** |  |  |
| Unknown (Idiopathic) | 18 (32.1%) | 5 (27.8%) |
| Stroke | 3 (5.4%) | 0 (0%) |
| Structural/Lesional | 35 (62.5%) | 13 (72.2%) |

In the full cohort of 56 participants, a total of 260 complete ARTiE Watch tests (198 interictal, 23 clinical events without electrographic correlates, and 39 ictal or postictal events with electrographic correlates) were administered during the study. Inclusion criteria for the final analyses were participants who had ARTiE Watch behavioral responsiveness assessments in both the interictal (baseline) periods (no ictal activity or clinical symptoms for one hour) and during electrographically confirmed seizure (ictal or postictal) periods.

In the 18 participants included in the final data analysis, there were 67 interictal (baseline) and 39 electrographic seizure (ictal-postictal) ARTiE Watch tests. Mean EEG seizure duration was 103 ± 72 s (mean ± SD) and mean clinical seizure duration was 167 ± 163 s. The first ARTiE Watch command prompt began a mean of 80 ± 76 s after EEG seizure onset and -27 ± 133 s before EEG seizure offset. ARTiE Watch testing onset occurred during the ictal phase in 23 (59%) seizures and in the postictal phase of 16 (41%) seizures (Table 4).

Seizures were also classified by EMU staff and clinical epileptologists according to ILAE guidelines (Fisher et al., 2017). Of the 18 participants, seizures were localized to the left hemisphere in 4 (22%), right hemisphere in 6 (33%), and bilateral hemispheres in 8 (44%) participants. Seizure onset involved the temporal lobe in 12 (67%), frontal lobe in 2 (11%), and was generalized or multifocal in 4 (22%) patients. Of the 39 seizures captured with ARTiE, there were 22 (56%) focal, 12 (31%) focal to bilateral tonic clonic, 1 (3%) generalized onset tonic-clonic, and 4 (10%) myoclonic seizures (Table 4). Of the 22 focal seizures, the EMU clinical staff classified 10 (45.5%) as focal aware seizures (FAS) and 12 (54.5%) as focal impaired awareness seizures (FIAS).

Assessment of interrater reliability comparing ARTiE Watch scoring utilizing vEEG for a subset of randomly 30 interictal and 30 seizure ARTiE Watch tests revealed a Cohen’s Kappa of 1.0.

### 3.2 Behavior is Impaired in Seizures Compared to Baseline Assessments

ARTiE Watch behavioral responsiveness was significantly decreased in participants undergoing testing during the ictal-postictal period of seizures compared to the baseline (interictal) period (Figure 3). We observed a significant decrease (p < 0.0001) in ARTiE watch testing scores in terms of Verbal Responsiveness, Motor Responsiveness, Memory Recall (largest magnitude of decrease), as well as in Overall Responsiveness in the seizure (ictal-postictal) period compared to baseline (interictal) testing (Figure 3 B – E).

**Figure 3.**
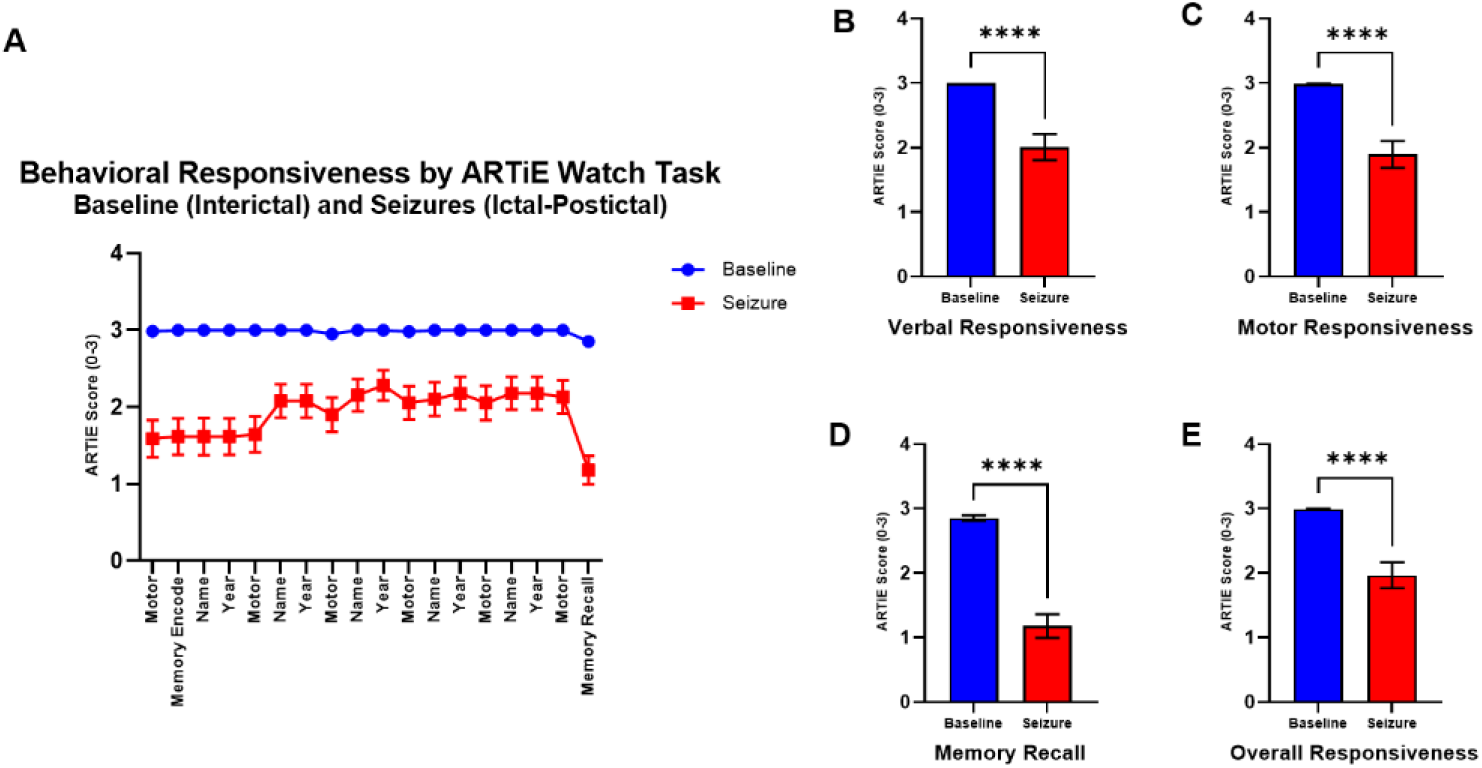
Baseline (interictal) and seizure (ictal-postictal) testing produce distinct ARTiE Watch behavioral responsiveness profiles. (a) There was greater impairment for each ARTiE Watch task during seizure (ictal-postictal) testing compared to baseline interictal testing. (b) Verbal responsiveness was impaired in the seizure (ictal-postictal) period. (c) Motor (nonverbal) Responsiveness was impaired in seizure (ictal-postictal) period. (d) Memory recall was lower in patients during seizures than baseline. (e) Overall behavioral responsiveness declined during seizure (ictal-postictal) ARTiE Watch testing compared to baseline (interictal) testing. Data are represented as the mean ± SEM. There were 18 participants who performed 67 baseline (interictal) tests and 39 seizure (ictal-postictal tests). Corrected for multiple comparisons, statistical significance is indicated as follows: \*\*\*\**p ≤ 0.0001*, \*\*\**p ≤0.001*, \*\**p ≤0.01*, \**p < 0.05*, *non-significant* (*p ≥ 0.05*).

### 3.3 Impairments Differ Between Focal, Tonic-Clonic (TC), and Myoclonic Seizures

ARTiE Watch behavioral responsiveness was severely impaired during TC seizures, variably impaired during focal seizures, and near baseline during myoclonic seizures (Figure 4). For all seizure types, there was a trend of improved behavioral responsiveness throughout the duration of the test excluding memory recall, which remained impaired at the conclusion of the testing sequence (Figure 4A). Tonic-clonic (focal and generalized onset) and focal seizures were significantly impaired on Verbal, Motor and Overall Responsiveness compared to baseline, but myoclonic seizures were not significantly impaired on these measures (Figure 4B, C, E). Tonic-clonic seizures were significantly more impaired than focal and myoclonic seizures on Verbal, Motor and Overall Responsiveness (Figure 4B, C, E). Memory Recall showed significant impairment compared to baseline for all seizure types including myoclonic seizures (Figure 4 D). Memory recall was significantly more impaired for tonic-clonic compared to focal seizures (Figure 4D).

**Figure 4.**
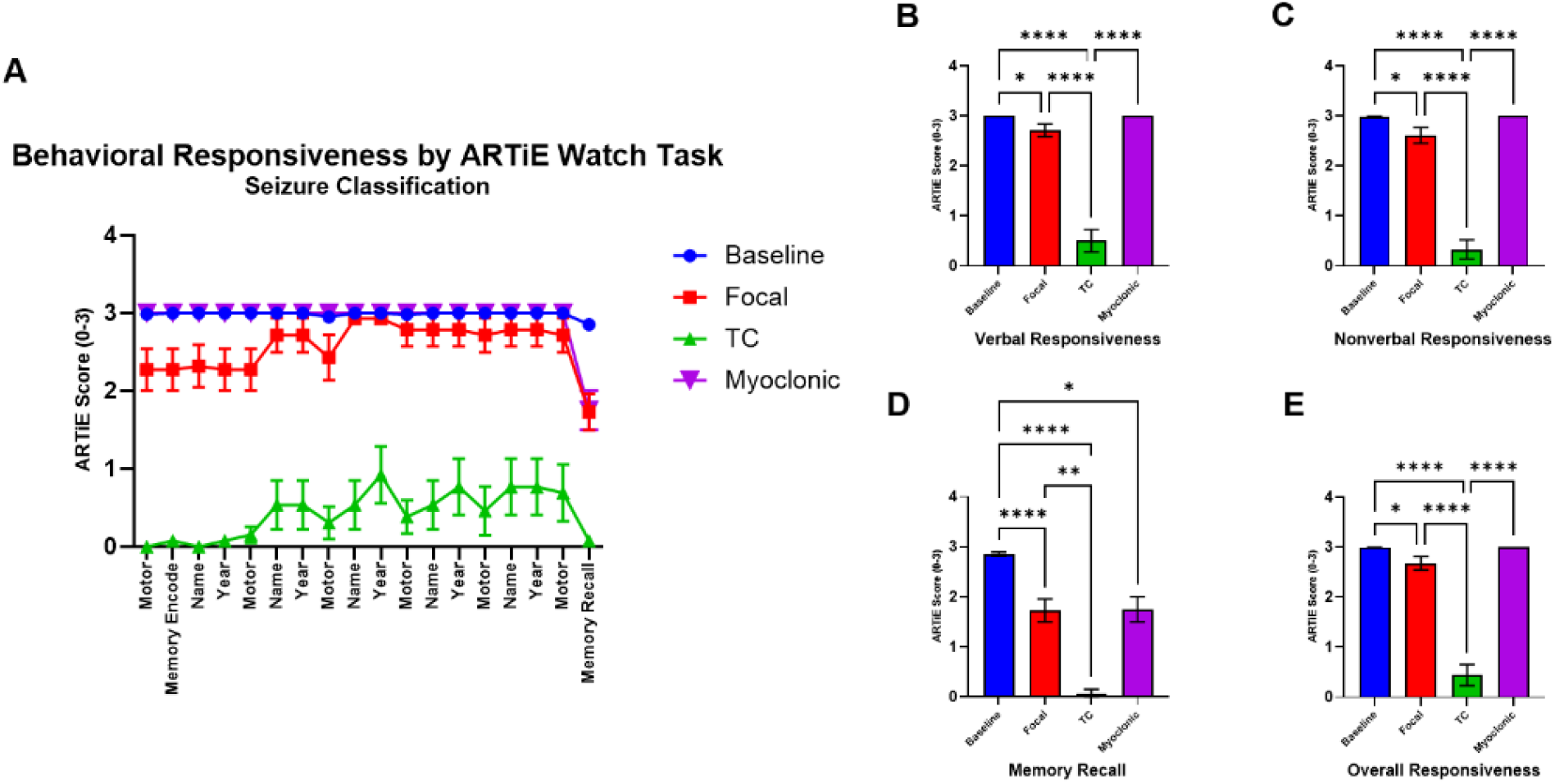
ARTiE Watch Behavioral Responsiveness Scores Differ Between Focal, Tonic-Clonic and Myoclonic Seizures. (a) Time course of impairment relative to onset of testing sequence show maximal impairment at early times for bilateral tonic-clonic (TC) and focal seizures, and minimal impairment with myoclonic seizures compared to baseline interictal testing. (b) Verbal Responsiveness was more severely impaired in TC compared to focal and myoclonic seizures. (c) Motor Responsiveness was more severely impaired in TC compared to focal and myoclonic seizures. (d) Memory Recall was impaired in all seizure types compared to baseline, and more severely impaired in TC compared to focal seizures(e) Overall Responsiveness was more severely impaired in TC compared to focal and myoclonic seizures. Data are represented as the mean ± SEM. There were 18 patients, 67 interictal tests, 4 myoclonic, 22 focal, and 13 tonic-clonic. The tonic-clonic category includes 12 focal to bilateral tonic-clonic seizures, and 1 generalized onset tonic-clonic seizure. Corrected for multiple comparisons, statistical significance is indicated as follows: \*\*\*\**p ≤ 0.0001*, \*\*\**p ≤0.001*, \*\**p ≤0.01*, \**p < 0.05*, non-significant (*p ≥ 0.05*).

### 3.4 Validation of ARTiE Watch Ratings by Standard Clinical Assessment

We compared ARTiE Watch behavioral ratings to standard clinical assessment by the epilepsy monitoring unit staff using the current ILAE classification (Fisher at al, 2017). There was greater impairment on ARTiE Watch behavioral responsiveness scores during focal impaired awareness seizures (FIAS) compared to focal aware seizures (FAS) (Figure 5). Verbal Responsiveness, Motor Responsiveness, and Overall Responsiveness were all significantly impaired in FIAS but not in FAS compared to baseline, and the same measures were significantly more impaired in FIAS than in FAS (Figure 5 B, C, E). However, delayed Memory Recall was significantly impaired in both FIAS and FAS (Figure 5 D).

**Figure 5.**
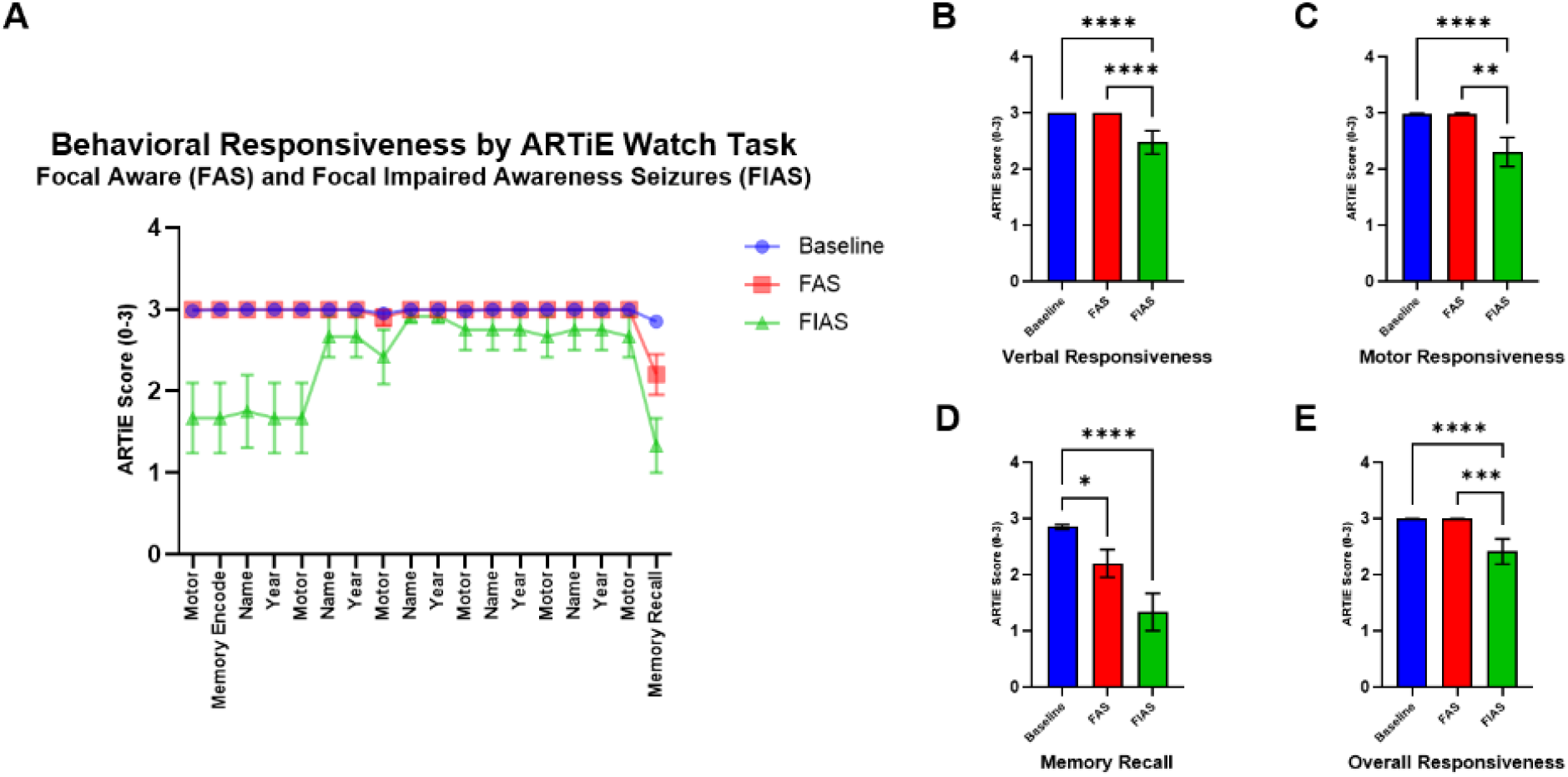
ARTiE Watch Behavioral Responsiveness Scores Compared to Standard Clinical Assessment. Focal seizures were labeled as focal aware seizures (FAS) or focal impaired awareness seizures (FIAS) by epilepsy monitoring unit clinical staff. (a) Time course of ARTiE Watch testing sequence shows impairment in FIAS, whereas FAS scores remain near baseline, except for delayed Memory Recall. Verbal Responsiveness on ARTiE watch testing was more severely impaired in FIAS compared to FAS. Motor responsiveness on ARTiE Watch testing was more severely impaired in FIAS compared to FAS. Memory Recall on ARTiE Watch testing was impaired in both FIAS and FAS compared to baseline. (e) Overall Responsiveness on ARTiE Watch testing was more severely impaired in FIAS compared to FAS. Data are represented as the mean ± SEM. There were 10 FAS and 12 FIAS evaluated. Corrected for multiple comparisons, statistical significance is indicated as follows: \*\*\*\**p ≤ 0.0001*, \*\*\**p ≤0.001*, \*\**p ≤0.01*, \**p < 0.05*, *non-significant* (*p ≥ 0.05*).

### 3.5 ARTiE Watch Ratings for Left, Right and Bilateral Focal Onset Temporal Lobe Seizures

For the 12 patients with temporal lobe seizure onset, there were 22 focal and – focal to bilateral tonic-clonic seizures. To investigate possible relationships between lobe of onset and behavioral impairment, we further classified the -- focal temporal lobe onset seizures into 11 left, 11 right and 4 bilateral onset seizures. Focal seizure onset locations were further classified by the clinical epileptology teams at each site using all available clinical information, including clinical history, video and scalp or intracranial EEG, neuroimaging, and surgical outcome if available. We found left, right and bilateral onset temporal lobe seizures led to impaired Verbal, Motor and Overall Responsiveness and well as impaired Memory Recall (Figure 6). Although bilateral onset seizures appeared to have a trend towards more severe impairment early in the testing sequence (Figure 6 A), we did not detect a significant difference in overall ARTiE Watch scores between left, right or bilateral onset temporal lobe seizures (Figure 6B-6E). However, all three seizure types showed significant differences from baseline during ARTiE Watch testing of Verbal Responsiveness, Motor Responsiveness, Memory Recall, and Overall Responsiveness (Figure 6B-6E).

**Figure 6.**
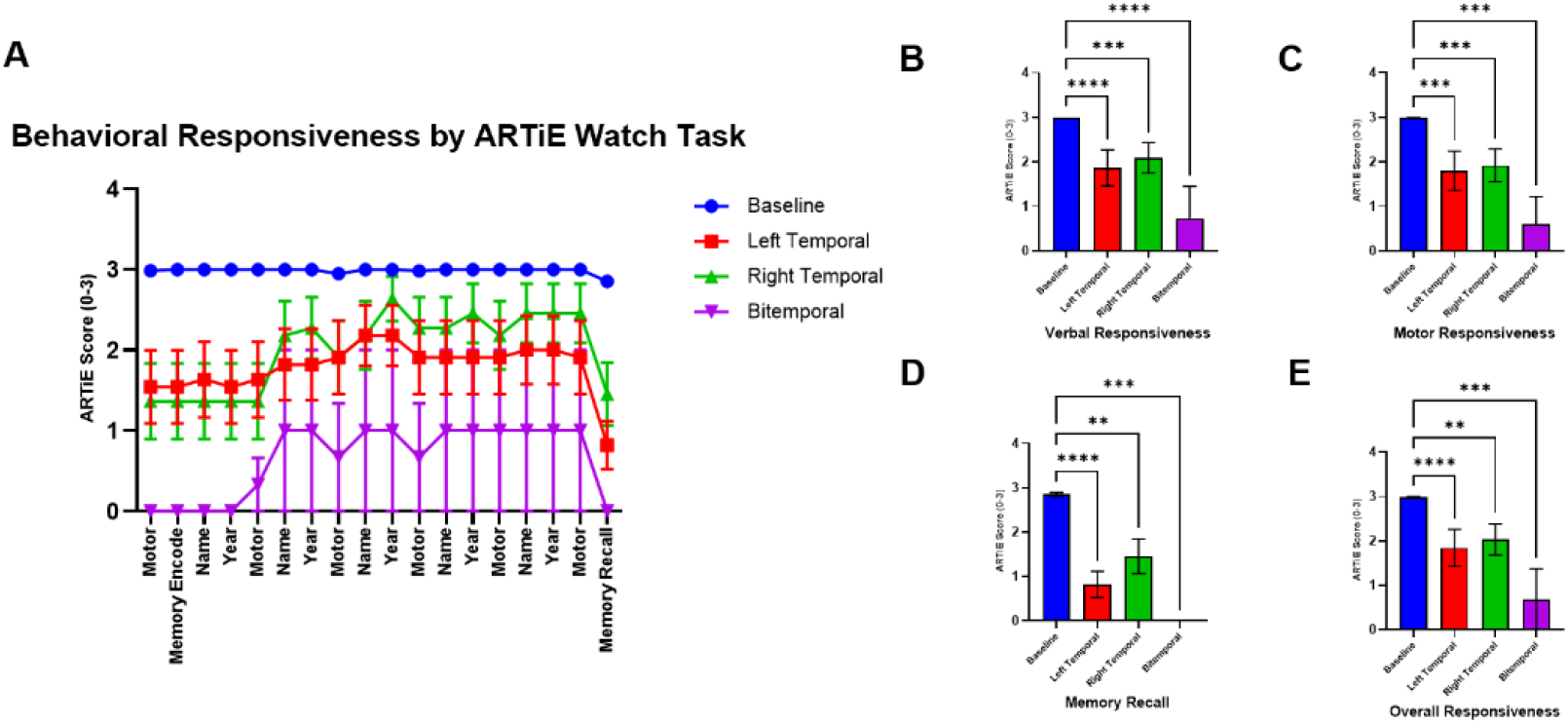
ARTiE Watch Behavioral Scores in Left, Right and Bilateral Onset Temporal Lobe Focal Seizures. (a) Left, right and bilateral onset temporal lobe seizures all caused impairment in ARTiE watch testing, with a trend towards more severe impairment with bilateral onset seizures. (b) Verbal Responsiveness was reduced compared to baseline, with no significant difference between seizure groups. (c) Motor responsiveness was reduced compared to baseline, with no significant difference between seizure groups. (d) Memory recall was impaired compared to baseline, with no significant difference between seizure groups. (e) Overall Responsiveness was reduced compared to baseline, with no significant difference between groups. Data are represented as the mean ± SEM. There were – left onset, -- right onset, and – bilateral onset temporal lobe seizures evaluated. Corrected for multiple comparisons, statistical significance is indicated as follows: \*\*\*\**p ≤ 0.0001*, \*\*\**p ≤0.001*, \*\**p ≤0.01*, \**p < 0.05*, *non-significant* (*p ≥ 0.05*).

## 4 DISCUSSION

In this study, we explored the feasibility and reliability of a mobile automated assessment of behavior using a cloud-based platform with a smartwatch and smartphone to automatically deliver standardized behavioral responsiveness assessments of patients in the EMU. The data show that the ARTiE Watch produced reliable, objective measures of behavioral impairment during the ictal and postictal periods in people with epilepsy compared to interictal baseline testing. We were able to assess behavioral responsiveness in multiple domains of function including orientation, motor responsiveness, verbal responsiveness, and memory recall.

ARTiE behavioral scores were analyzed in relation to seizure type, clinical seizure classification, and temporal lobe seizure onset. The greatest ARTiE impairment occurred during tonic-clonic seizures, followed by mixed impairment during focal seizures, and minimal impairment during myoclonic seizures. Notably, the time lag between seizure onset and ARTiE testing likely hindered the evaluation of brief myoclonic seizures. The more severe impairment during tonic-clonic compared to focal seizures with ARTiE Watch behavioral testing is consistent with previous studies using human-administered behavioral testing, which also showed more severe impairment in tonic-clonic seizures (McPherson et al., 2012).

ARTiE Watch behavioral scores were in agreement with clinician assessments, showing more severely impaired ARTiE Watch responsiveness scores for FIAS compared to FAS. It should be noted that clinicians viewing videos of patients during these seizures were able to observe the ARTiE Watch behavioral testing, so the two forms of assessment were not truly independent. Nevertheless, the results demonstrate that quantitative ARTiE Watch testing scores were in agreement with more informal clinically-based evaluations. Interestingly, the memory recall scores were impaired in both seizures classified as FIAS and FAS based on clinical assessment, indicating that recall of events during seizures may be less reliable than responsiveness testing during seizures for classification of FIAS versus FAS. These findings add to the growing literature suggesting that responsiveness should be formally included in the definition of FIAS versus FAS in addition to memory recall (Contreras Ramirez et al, 2022a, b).

ARTiE Watch testing scores did not reveal a statistically significant difference between behavioral impairment in temporal lobe seizures with left, right or bilateral onset. There did appear to be trend towards greater impairment with bilateral onset temporal lobe onset seizures, so it will be important to investigate this further in a larger sample size. Of note, previous work with intracranial EEG suggests that although both left and right onset temporal lobe seizures can cause impaired consciousness, impairment is more commonly observed in left onset seizures (Englot et al, 2010), so this question should also be investigated further in a larger patient sample.

These results add to the literature on standardized behavioral responsiveness testing during seizures in the EMU (Yang et al., 2012; Bauerschmidt et al., 2013; Cunningham et al., 2014; Touloumes et al. 2016; Benickzy et al, 2016), but here is extended to a mobile platform suitable for deployment in ambulatory subjects living in their natural home environment. Such a platform may have wide application if family members can activate the behavioral testing, or with home ambulatory video/EEG systems which may have capability for automatic seizure detection. In another important application, combination of the ARTiE Watch with implanted devices capable of automatic seizure detection and neurostimulation can serve as a highly useful way to objectively evaluate behavior during seizures in the home setting, to assess device therapeutic efficacy. This approach is being used in a clinical trial currently underway (Yadav et al., 2022).

### 4.2 Limitations and Future Directions

The study is an exploratory pilot study in a relatively modest number of participants. A larger representative sample of people with epilepsy should be explored in the future. The time between EEG seizure onset and ARTiE initiation is limited here by the manual initiation and is inadequate for short seizures (e.g. myoclonic seizures). The interpretation of responsiveness is human dependent and thus susceptible to bias, although the interobserver reliability was excellent.

During EMU evaluation, patients often undergo seizure induction measures (e.g. tapering anti-seizure medications, hyperventilation, sleep deprivation) which may extend the duration, severity, and morphology of the peri-ictal, ictal, and postictal phases. In future research, we plan to expand the study to ambulatory participants implanted with brain sensing devices that can detect pathological interictal and ictal discharges that initiate ARTiE behavioral testing. The application to ambulatory participants living in their natural home environment will enable capturing behavioral phenotypes of a broad range of electrophysiological patterns in ecologically valid environments (Kremen et al., 2018; Kremen et al. 2024).

## 5 CONCLUSIONS

This is the first multicenter EMU study using a novel behavioral responsiveness and electrophysiology platform for standardized mobile behavioral responsiveness testing during seizures. Our results indicate that the ARTiE Watch deployed utilizing a wearable and mobile health platform is feasible, and can provide reliable, objective measures of behavioral impairment across multiple functional domains. The ARTiE Watch system can offer clinicians an automated objective quantitative tool to different behavioral responsiveness during, between, and after seizures. While our system was motivated by epilepsy, the ARTiE Watch system can be used to study other neurological and psychiatric disorders.

## Data Availability

All data produced in the present study are available upon reasonable request to the authors.

## AUTHOR CONTRIBUTIONS

LW, ADL, VK, KM, VS, CB, JG, GW, HB and the START Project Collaborative Team contributed to the study’s conception and design. LW, CM, GO, TY, DC, VK, GW, HB contributed to the acquisition and analysis of data. LW, TY, VK, GW, HB contributed to drafting a significant portion of the manuscript or figures.

## ACKNOWLEDGMENTS

This study was supported by NIH UG3/UH3 NS112826. We are grateful to the following for technical, administrative or regulatory support for this project: Abbey Becker, Amy Hummel, David Linde, Delana Weis, Irina Korytov, Jon Giftakis, Kathy Polak, Manichanh Howell, Robert Raike, Samuel Heck, Starr Guzman, Thaddeus Brink, Tyler Hamilton, Zi Kai (Kyle) Soo.

## APPENDIX START (Stimulation of the Thalamus for Arousal Restoral in Temporal lobe epilepsy) Project Collaborative Team

Abhijeet Gummadavelli, Allyson Derry, Anastasia Kanishcheva, Angela Waszkiewicz, Barbara Jobst, Benjamin Brinkmann, Bogdan Patedakis Litvinov, Brian Lundstrom, Brian Rutt, Catherine Doucet, Charlotte Jeffreys, Christopher Benjamin, Christopher R. Butson, Courtney Yotter, Dennis Spencer, Devon Cormier, Eric B. Geller, Eun Young Choi, Eva Alden, Eyiyemisi Damisah, George Thomas, George W. Culler, Grant Moncrief, Gregory Worrell, Hal Blumenfeld, Imran Quraishi, Jaimie Henderson, Jamie Van Gompel, Jason Gerrard, Jennifer Hong, Jonathan Lee Baker, Joseph Giacino, Joshua Aronson, Karla Crockett, Kate Christison-Lagay, Kimberly Bailey, Kristine DaCosta, Krzysztof A. Bujarski, Kyle O’Sullivan, Lawrence Hirsch, Lydia Wheeler, Matthew Hook, Maxime Oriol, Nicholas D. Schiff, Nicholas Gregg, Patrice Lauture, Paul E.Croarkin, Robert Roth, Ruben I. Kuzniecky, Stephen Meisenhelter, Steven Messina, Taruna Yadav, Vaclav Kremen, Vladimir Sladky, Yinchen Song, Zan Ahmad, Zheng Zhang.

